# Maternal sleep during the first ten years of the child’s life

**DOI:** 10.1101/2025.07.15.25331584

**Authors:** Natalia Schwarzkopf, Mayda Rivas, Florencia Peña, Andrea Devera, Pablo Torterolo, Ana Hernández, Luciana Benedetto

## Abstract

Main sleep disturbances during motherhood occur during the first weeks after childbirth, but sleep continues to be affected until the baby sleeps through the night. However, most studies focus their attention up to 18 months after delivery. It is our interest to determine sleep quality of mothers with children from 0 to 10 years sleep, and the sociocultural factors associated with it. We designed a cross-sectional quantitative study consisting of an anonymous survey online that included the Pittsburgh Sleep Quality Index (PSQI), sociodemographic data and nocturnal family dynamic questions. The main findings reveal that the quality of maternal sleep was poor for all mothers regardless of the child’s age. Specifically, during the first two years after childbirth maternal sleep quality was particularly low, with a significant improvement towards age 3, worsening again at the ages of 4 and 5 of the child, to finally improve towards the ages of 8 to 10. Additionally, higher levels of education and performing physical activity increased the probability of having good sleep quality. Conversely, child’s awakenings increased the odds for the mother of having poor sleep quality at all stages of the child. For mothers with younger children, additional aspects of their children’s sleep and social and nocturnal family dynamic also predict their sleep quality. The current results expose that maternal sleep disturbances extend far beyond the postpartum period. To the best of our knowledge, this is the first report that focuses on maternal sleep until the age of 10 years of the child.

## Introduction

In all mammalian mothers studied, sleep alterations occur at some point of the postpartum period [1–6]. In human mothers, sleep disturbances, such as fragmentation, have been consistently reported during the postpartum period [1, 7–11]. In addition, although the main sleep disruptions occur during the first weeks after delivery and may persist for up to six months, full sleep satisfaction is not typically regained at least until the child is at least six years old [12–15].

Sleep is influenced by a complex interplay between psychosocial, physiological, and environmental factors [16]. During the postpartum period, the mother’s age, parity, culture, and social environment impact sleep [16–18]. However, the primary sleep disturbances during early motherhood are mainly caused by the polycyclic sleep pattern of the newborn [1]. Although most babies achieve sleep consolidation by the age of 4-12 months (often referred to as “sleeping through the night), not all children reach this milestone by that age [19–21]. In addition, approximately 20% of preschool and school-aged children experience various sleep disturbances [22–30]. These disturbances can, in turn, negatively impact the mother’s ability to sleep uninterrupted at night [31].

As sleep characteristics vary depending on the sociocultural factors, it is not surprising that different aspects of sleep, such as duration and time onset, vary across different countries [32]. However, most research has been focused on high socioeconomic countries [32, 33], and a meta-analysis found high variability in sleep characteristics in low-middle income countries and highlights the need for more epidemiological research about basic sleep health parameters in these countries [33]. In particular, studies regarding maternal sleep are not an exception to this matter, and there are almost no studies focusing on maternal sleep in South American countries, with only one article published from Brazil [34]. In addition, despite a large amount of literature on how human mothers sleep during the first months after delivery in various countries around the world [35], the literature about maternal sleep beyond this stage is scarce [15].

The current report is a cross-sectional study, where we describe the subjective sleep quality of women with children from birth to 10 years of life from Uruguay, using the Pittsburgh Sleep Quality Index (PSQI), a survey extensively used across nations for general population [33], and also mothers [36]. In addition, we analyze possible factors that may improve or worsen maternal sleep quality at different stages of the child, including socio-demographic variables (maternal age, education level, employed), infant sleep characteristics, and nocturnal family dynamic. To our knowledge, this is the first report about maternal sleep in a South American country beyond the first months of the postpartum period, highlighting the significance of our data.

## Methods

### Participants

A total of 1189 mothers of young children from 0 to 10 years old participated in this study. No exclusion criteria were applied to either the mothers or the children in order to include the entire population [37].

### Study design

This study was a cross-sectional quantitative design using an anonymous online survey related to most usual habits and practices of sleep over the last month. The information analyzed in the present article is part of a larger project aimed at exploring the link between maternal sleep, nocturnal family dynamic and sociocultural variables.

### Protocol

This study was conducted according to the principles of the Declaration of Helsinki and approved by the Ethics Committee of the School of Medicine, Universidad de la República, Uruguay (N° 070153-000023-22).

All data was collected from an online survey from December 2022 to June 2023, where the link to complete the survey was available or sent, together with the general information of the project. Social networks (Twitter, Instagram and Facebook) and a list of personal contacts were used to invite mothers to complete it.

The first item in the survey was to read any possible cons and benefits about the completion of the survey and then approve the informed consent.

Completion of the survey was voluntary, having the possibility to finish the questionnaire at any time. No identifying information was collected.

### Measures

#### General information about the mother and the child

A first set of questions related to sociodemographic information from both the child and the mother (age, work, education) and family composition was asked. Afterwards, we asked questions regarding the type of feeding and child’s gender. Mothers with more than one child were asked to complete the questionnaire based on the child who had the greatest impact on their sleep pattern.

#### Nighttime sleep family dynamic

A series of queries were aimed at knowing about nocturnal family dynamic during the last month. These included the child’s sleeping arrangement, the number of nighttime awakenings experienced by both the mother and the child, the parentś attitude towards the child’s awakenings, whether other adults assisted during these awakenings, how the child returned to sleep and how long it took.

#### Subjective sleep quality

The final questions were the PSQI, that explores sleep quality related to the last month. This questionnaire was originally developed by [38] adapted to Spanish [39], and previously demonstrated to obtain reliable data in postpartum women [14, 40].

### Data analysis

Descriptive analysis was carried out to describe demographic characteristics, as well as the PSQI and its dimensions.

The PSQI consists of 19 self-rated questions and five questions to be answered by bedmates or roommates [38]. The latter questions are not used for analysis. The questions generate seven scores (with subscales ranged 0–3): sleep quality, sleep latency, sleep duration, habitual sleep efficiency, sleep disturbance, use of sleeping medication, and daytime dysfunction. The sum of these components scores generates one general score ranged from 0 to 21, where higher scores represent poorer subjective sleep quality, and scores > 5 indicate poor sleep quality (sensitivity 89.6%, specificity 86.5%) [38].

For the first descriptive analysis of how sleep quality varies according to the childreńs age, the mothers were grouped into 9 categories according to the following ages of their child: 0-5 months (n = 154), 6-11 months (n = 162), 12-17 months (n = 152), 18-23 months (n = 106), 2 years (n = 152), 3 years (n = 102), 4-5 years (n = 138), 6-7 years (n = 114), 8-10 years (n = 109). For further analysis, to facilitate the interpretation, we re-grouped the mothers into three categories according to the childreńs age: a) 0-2 years, infant-toddler; 3-5 years, preschool; 6-10 years, school.

### Statistical analysis

We used Kolmogorov-Smirnov normality test to verify the normality of the continuous variables used. Since data did not follow a normal distribution, results are presented as medians (Q1, Q3) and mean ranks. In order to statistically analyze group differences on global PSQI and its components, we used the Kruskal–Wallis test followed by Dunn’s *post hoc* test.

To analyze the possible predictors for global PSQI score we used multinomial logistic regression models similar to [41–43]. For that purpose, as this variable did not follow a normal distribution, we categorized the variable into 4 categories: PSQI 0-5 (PSQI_0-5_), PSQI 6-9 (PSQI_6-9_), PSQI 10-13 (PSQI_10-13_), PSQI 14-21 (PSQI_14-21_). Independent variables were: motheŕs age, being employed/unemployed, education, biological sex of the child, family composition, sleeping arrangements, collaboration of other adults during nocturnal care of the child, child’s nocturnal awakening, performing naps and physical activity.

For all analysis, the null hypothesis was rejected when p < 0.05.

## Results

### Sample characteristics

A total of 1.189 mothers completed the survey, with no participants excluded from the analysis. Among the participants, 13 mothers reported symptoms of depression, 4 had anxiety syndrome, 6 children had specific genetic syndromes, and 6 children were diagnosed with autism spectrum disorder (ASD).

Demographic characteristics of the sample are presented in Table 1. The median value for the age of the mothers was 37 years (Q1 = 33; Q3 = 40), with significant differences among all groups: infant-toddler (35, 32-38; median, Q1-Q3), preschool (38, 34-40) and school (40, 37-43); all *p* values of Dunńs test were < 0.0001. The median number of infants in the families was one (Q1 = 1; Q3 = 2). The majority of participants had completed college education (69.2%), lived in nuclear families (families composed of both parents and the child; 90%), and were employed (90%).

**Table 1.** Socio-demographic data of all mothers.

| **Demographic variables** |  |
| --- | --- |
|  | **Mdn (Q1-Q3)** |
| **Maternal age (years)** |  |
|  | 37 (33-40) |
| **Number of children** |  |
|  | 1 (1-2) |
|  | **n (%)** |
| **Children gender** |  |
| Female | 591 (49.7) |
| Male | 598 (50.3) |
| **Highest education level completed** |  |
| Elementary school | 40 (3.4) |
| High school | 326 (27.4) |
| College | 823(69.2) |
| **Employment status** |  |
| Full or part time | 1081 (91) |
| No employed | 108 (9) |
| **Working shifts** |  |
| Day shift | 890 (82.3) |
| Night shift | 12 (1.1) |
| Shift work | 179 (16.6) |
| **Family composition** |  |
| Nuclear family | 1070 (90) |
| Single parent family | 104 (8.7) |
| Extended family | 15 (1.3) |

Data are presented as number and percentage (n, %) or median (Mdn) and quartiles 1 and 3 (Q1-Q3).

### General sleep characteristics

The median PSQI score for all mothers was 9 (Q1 = 6, Q3 = 12), with 85% of participants scoring above the PSQI cut-off score of 5. In Figure 1, it is depicted how the PSQI varies along the first 10 years of life. When comparing the global PSQI at the different ages of the children with the one in the first group (mothers with children from 0-5 months), we found statistical differences between this group and the groups of mothers with children aged 2 years and 8 to 10 years; global PSQI values in the other groups did not reach statistical significance.

**Figure 1.**
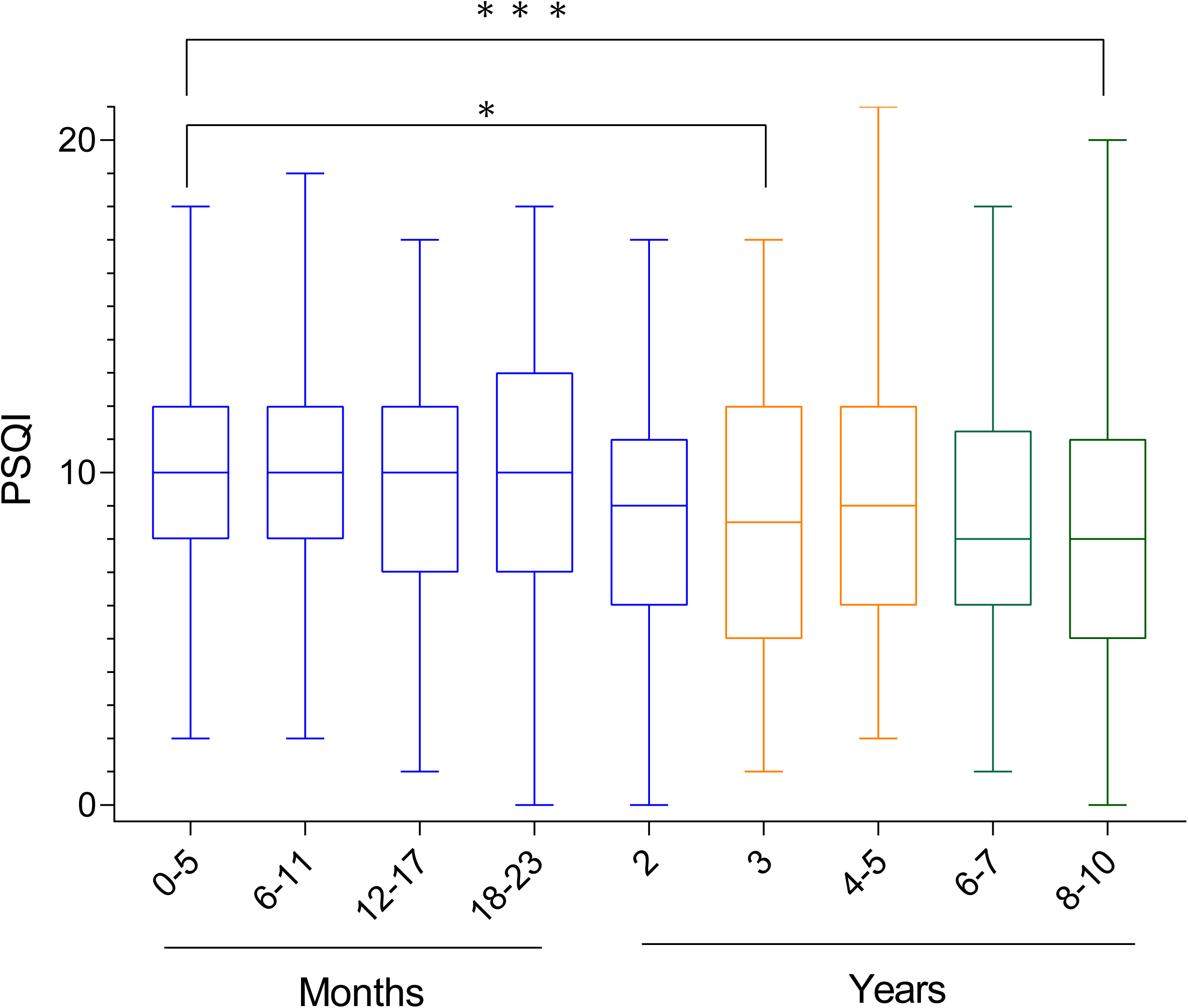
Global sleep quality scores for mothers across different ages of their children. Different colors indicate the regrouping performed for posterior analysis of possible predictors of PSQI values. Statistical comparisons were made using ANOVA and Dunńs test, with mothers of 0–5-month-old infants as the reference group. *, *p* < .005; ***, p < .0001.

As seen in Table 2, when comparing the global PSQI among mother with infant-toddler, preschool, and school children, we found that the PSQI of mothers with infant-toddler children was significantly higher compared to that of preschool and school children (p = <.001). Specifically, mothers with infant-toddler children had worse sleep quality compared to both mothers with preschool and school children, but there was no statistical difference between these two latter groups. In addition, 85.8 % of the mothers in the infant-toddler group had sleep disturbances, while 80 % in the preschool group and 72.6 % in the school group presented a PSQI > 5.

**Table 2.** PSQI components. Mothers were divided into three groups according to following ages of the child: infant-toddler (0-2 years), pre-school children (3-5 years), school children (6-10 years).

| Variable | Infant-toddler<br>(n=726) | | Preschool<br>(n=240) | | School<br>(n=223) | | $\frac{P}{K-W}$ | Post Hoc | | |
| --- | --- | --- | --- | --- | --- | --- | --- | --- | --- | --- |
|  | Mdn (Q1-Q3) | MR | Mdn (Q1-Q3) | MR | Mdn (Q1-Q3) | MR |  | + | # | * |
| Global sleep quality | 10 (7-12) | 637.1 | 8 (6-12) | 555.0 | 8 (5-11) | 501.1 | < 0.001 | 0.001 | < 0.001 | 0.271 |
| Subjective sleep quality | 2 (1-2) | 632.2 | 2 (1-2) | 547.2 | 1 (1-2) | 525.0 | < 0.001 | 0.001 | < 0.001 | 1.001 |
| Sleep latency | 1 (0-2) | 590.1 | 2 (0-2) | 631.4 | 1 (0-2) | 569.1 | 0.109 | - | - | - |
| Sleep duration | 1 (1-2) | 633.3 | 1 (1-2) | 553.5 | 1 (1-1) | 513.3 | < 0.001 | < 0.002 | < 0.001 | 0.503 |
| Sleep efficiency | 2 (1-3). | 670.5 | 1 (0-2). | 509.6 | 1 (0-2) | 451.1 | < 0.001 | < 0.001 | < 0.001 | 0.031 |
| Sleep disturbance | 1 (1-2). | 605.6 | 1 (1-2) | 609.6 | 1 (1-2) | 543.6 | < 0.001 | 1.001 | 0.017 | 0.047 |
| Sleep medication | 0 (0-0) | 562.0 | 0 (0-0) | 623.5 | 0 (0-1) | 664.8 | < 0.001 | 0.001 | < 0.001 | 0.079 |
| Daytime dysfunction | 2 (1-2) | 616.6 | 2 (1-2) | 593.0 | 1 (1-2) | 527.5 | 0.001 | 0.982 | < 0.001 | 0.086 |

Values are presented as median (Mdn) and quartiles 1 and 3 (Q1–Q3) and mean rank (MR). Higher values indicate poorer sleep quality. Significant differences among groups were analyzed using Kruskal Wallis and Dunn’s test as *post hoc*. Plus symbol (+) indicates a significant difference between mothers of infants-toddlers and preschoolers; numeral symbol (#) indicates a significant difference between mothers of infants-toddlers and school children; asterisk symbol (\*) indicates a significant difference between mothers of preschoolers and school children.

Regarding the components of PSQI, our results indicate statistically significant differences in all components of PSQI except for sleep latency when comparing mothers of infant-toddler children with mothers of school children or both preschool and school children (Table 2). Specifically, statistical tests revealed that mothers of infant-toddler children had poorer subjective sleep quality scores, shorter sleep durations, lower sleep efficiency and consumed less sleep medication when compared to mothers of both preschool and school children. Additionally, the sleep disturbance and daytime dysfunction were higher in mothers of infant-toddlers compared to mothers of schoolers. When comparing PSQI components of mothers of preschoolers and schoolers, sleep efficiency was reduced, and sleep disturbances were higher in the former mothers.

### Sleep quality predictors

To identify which variables could predict bad sleep quality in each group of mothers, the PSQI was divided into four categories: PSQI_0-5_, PSQI_6-9_, PSQI_10-13_, PSQI_14-21_. For this purpose, the category considered good sleep quality (PSQI<5) was taken as the reference to compare with the other categories.

#### Mothers with infant-toddler children (ranged from 0-2 years of age)

The distribution of mothers with infant-toddlers according to the PSQI categories was: PSQI_0-5_, 14.3%; PSQI_6–9_, 35.4%; PSQI_10–13_, 37.9%; PSQI_14-21_, 12.4%.

Regarding the main socio-behavioral factors of this group, in 41.5% of cases, the mother was the primary caregiver during nighttime awakenings. In addition, 46.6% reported never taking naps and 56.3% of mothers reported engaging in less than 30 minutes of physical activity per day. In terms of sleep arrangements, the most prevalent was bed-sharing, reported by 42.8% of participants.

All the variables that were statistically significant or tended to be significant as predictors of the PSQI are detailed in Table 3 (complete data is available in supplementary data, Table S1).

**Table 3.**
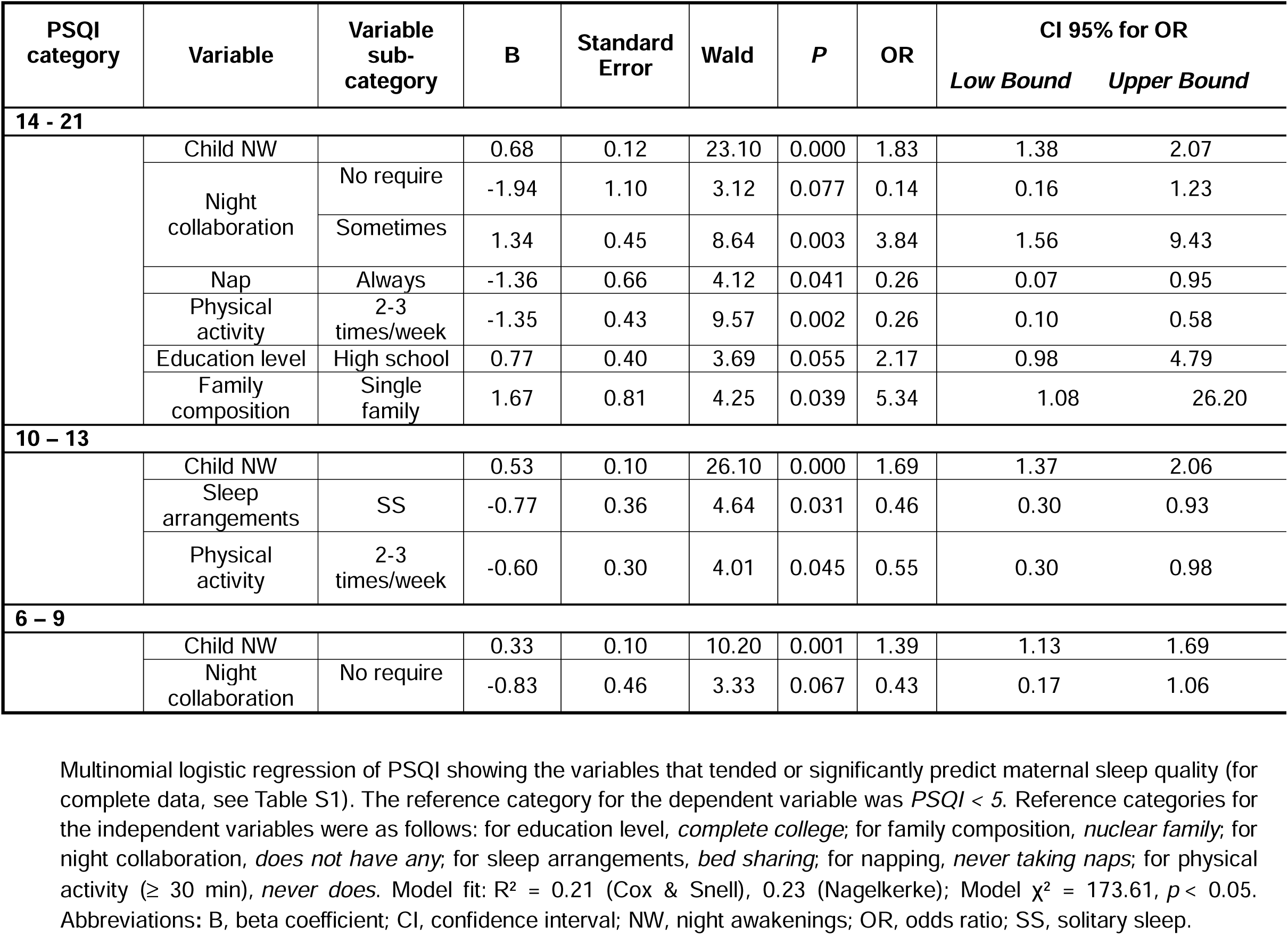
Main variables that predict maternal sleep quality in the group of mothers with infant-toddler children (0-2 years).

| PSQI category | Variable | Variable sub-category | B | Standard Error | Wald | P | OR | CI 95% for OR |  |
| --- | --- | --- | --- | --- | --- | --- | --- | --- | --- |
|  |  |  |  |  |  |  |  | Low Bound | Upper Bound |
| 14 - 21 |  |  |  |  |  |  |  |  |  |
|  | Child NW |  | 0.68 | 0.12 | 23.10 | 0.000 | 1.83 | 1.38 | 2.07 |
|  | Night collaboration | No require | -1.94 | 1.10 | 3.12 | 0.077 | 0.14 | 0.16 | 1.23 |
|  |  | Sometimes | 1.34 | 0.45 | 8.64 | 0.003 | 3.84 | 1.56 | 9.43 |
|  | Nap | Always | -1.36 | 0.66 | 4.12 | 0.041 | 0.26 | 0.07 | 0.95 |
|  | Physical activity | 2-3 times/week | -1.35 | 0.43 | 9.57 | 0.002 | 0.26 | 0.10 | 0.58 |
|  | Education level | High school | 0.77 | 0.40 | 3.69 | 0.055 | 2.17 | 0.98 | 4.79 |
|  | Family composition | Single family | 1.67 | 0.81 | 4.25 | 0.039 | 5.34 | 1.08 | 26.20 |
| 10 – 13 |  |  |  |  |  |  |  |  |  |
|  | Child NW |  | 0.53 | 0.10 | 26.10 | 0.000 | 1.69 | 1.37 | 2.06 |
|  | Sleep arrangements | SS | -0.77 | 0.36 | 4.64 | 0.031 | 0.46 | 0.30 | 0.93 |
|  | Physical activity | 2-3 times/week | -0.60 | 0.30 | 4.01 | 0.045 | 0.55 | 0.30 | 0.98 |
| 6 – 9 |  |  |  |  |  |  |  |  |  |
|  | Child NW |  | 0.33 | 0.10 | 10.20 | 0.001 | 1.39 | 1.13 | 1.69 |
|  | Night collaboration | No require | -0.83 | 0.46 | 3.33 | 0.067 | 0.43 | 0.17 | 1.06 |

A predictor for bad sleep for all categories of PSQI was the child’s awakenings during the night. Also, a single-parent family composition significantly increased the odds of being in the PSQI_14-21_ category by almost 5 times. The level of education tended to impact on the motheŕs sleep quality. Specifically, mothers who only completed high school had 2 times more probability of having poor quality of sleep (PSQI_14-21_) compared to mothers with complete college education.

Various factors increase the probability of good sleep quality. Specifically, a mother whose child sleeps alone significantly increased (PSQI_10-13_) the probability of having good quality of sleep compared to a mother that co-sleeps with the child. Also, physical activity for at least 30 minutes two or three times a week is also a predictor of good sleep quality compared to a mother who did not engage in any physical activity for the categories PSQI_10-13_ and PSQI_14-21_. Taking naps always or almost always, in the category PSQI_14-21_, also predicted better sleep quality. If the child did not require attention during the night (PSQI_14-21,_ PSQI_6-9_), the mothers tended to have a better quality of sleep. In opposition, if their partner collaborated sometimes during the child’s awakenings, it increased the probability of being in the category PSQI_14-21_ by almost 4 times.

#### Mothers with preschool children (ranged from 3-5 years of age)

The category distribution of the PSQI in this group was: PSQI_0-5_, 19.7 %; PSQI_6–9_,31.0 %; PSQI_10–13_; 40.1 % PSQI_14-21_,9.2 %.

Regarding maternal socio-behavioral factors, in 34.7% of cases, the mother was the primary caregiver during nighttime awakenings of the child. Additionally, 64.9% indicate never taking naps and 44.4% of participants reported engaging in less than 30 minutes of physical activity per day. Concerning sleep arrangements, bed-sharing was reported in 29.3% of cases.

Main significant results from multivariable analysis are depicted in Table 4 (all data are detailed in Table S2). Specifically, physical activity for at least 30 minutes two to three times per week was a significant predictor of good sleep quality among mothers in the PSQI_10–13_ category, compared to those who reported no physical activity. A similar trend was observed for the PSQI_6–9_ and PSQI_14–21_ categories, although it did not reach statistical significance.

**Table 4.** Main variables that predict maternal sleep quality in the group of mothers with preschool children (3-5 years).

| PSQI category | Variable | Variable sub-category | B | Standard Error | Wald | P | OR | CI 95% for OR<br>*Low Bound Upper Bound* |  |
| --- | --- | --- | --- | --- | --- | --- | --- | --- | --- |
| 14 - 21 |  |  |  |  |  |  |  |  |  |
|  | Child NW |  | 1.12 | 0.34 | 12.30 | 0.000 | 3.32 | 1.72 | 6.44 |
|  | Sleep arrangements | SS | -1.48 | 0.71 | 4.30 | 0.038 | 0.22 | 0.06 | 0.92 |
|  | Physical activity | 2-3 times/week | -1.50 | 0.85 | 3.19 | 0.074 | 0.22 | 0.04 | 1.15 |
| 10 – 13 |  |  |  |  |  |  |  |  |  |
|  | Child NW |  | 0.55 | 0.22 | 5.83 | 0.016 | 1.73 | 1.11 | 2.71 |
|  | Sleep arrangements | SS | -1.48 | 0.52 | 7.76 | 0.005 | 0.23 | 0.08 | 0.66 |
|  |  | RS | -2.07 | 0.78 | 6.96 | 0.008 | 0.12 | 0.03 | 0.57 |
|  | Physical activity | 2-3 times/week | -1.13 | 0.48 | 5.54 | 0.019 | 0.32 | 0.12 | 0.82 |
|  | Work | Unemployed | -1.31 | 0.73 | 3.21 | 0.073 | 0.26 | 0.09 | 4.56 |
| 6 – 9 |  |  |  |  |  |  |  |  |  |
|  | Mother’s age |  | 0.13 | 0.04 | 7.91 | 0.005 | 1.14 | 0.80 | 2.37 |
|  | Education level | School | 2.16 | 1.27 | 2.86 | 0.091 | 8.69 | 0.70 | 106.72 |
|  | Physical activity | 2-3 times/week | -0.89 | 0.49 | 3.31 | 0.069 | 0.41 | 0.16 | 1.07 |

The number of child’s awakenings was associated with an increase in the probability of having poor sleep quality, increasing by 3.3 times the odds of being in the PSQI_14-21_ category, and 1.7 times in the PSQI_10-13_ category, with no significant effect observed in the PSQI_6-9_ group. Conversely, mothers whose children slept in a separate room or shared the room but on a separate surface had approximately 4-5 times more probability of having good sleep quality compared to those who co-sleep, particularly within the PSQI_14-21_ and PSQI_10-13_ categories.

Another predictor of poor sleep quality was the motheŕs age, which increased the probability of being in the category of PSQI_6-9_ instead of the good sleep category.

Finally, unemployment tended to increase the probability of having a better sleep quality (PSQI_10-13_), while lower levels of education (PSQI_6-9_) tended to increase the probability of worsening sleep quality.

#### Mothers with school children (ranged from 6-10 years of age)

The percentage of mothers in each PSQI category in this group was: PSQI_0-5_, 28.7 %; PSQI_6–9_, 28.3 %; PSQI_10–13_; 33.1 % PSQI_14-21_, 9.9 %.

In relation to socio-behavioral factors, in 28.7% of cases, the mother was identified as the primary caregiver during nighttime awakenings. Besides, 67.3% indicate never taking naps and 39.0% of participants reported engaging in less than 30 minutes of physical activity per day. With respect to sleeping arrangements, bed-sharing was reported in 17.5% of cases.

The variables that had a tendency or significantly predict the probability to be in certain PSQI category are shown in Table 5; complete data are available in Table S3. The level of education, performing physical activity and child’s awakenings impact on the quality of sleep. Specifically, higher level of education significantly predicted a better sleep quality in all categories (PSQI_6-9,10-13,14-21_). Practicing physical activity at least two to three times per week also significantly increased (PSQI_6-9_) or tended to increase (PSQI_10-13, 14-21_) the probability of having a PQSI < 5, similar to practicing physical activity four or more times a week. In addition, older mothers had a higher probability of being in the category PSQI_0-5_ compared to the category PSQI_10-13_. Taking naps almost always (PSQI_14-21_, PSQI_10-13_) significantly predicted better sleep quality.

**Table 5.** Main variables that predict maternal sleep quality in the group of mothers with school children (6-10 years).

| PSQI category | Variable | Category | B | Standard Error | Wald | P | OR | CI 95% for OR |  |
| --- | --- | --- | --- | --- | --- | --- | --- | --- | --- |
|  |  |  |  |  |  |  |  | Low Bound | Upper Bound |
| 14 - 21 |  |  |  |  |  |  |  |  |  |
|  | Child NW |  | 1.08 | 0.40 | 7.15 | 0.008 | 2.94 | 1.33 | 6.50 |
|  | Nap | Sometimes | -1.62 | 0.82 | 3.87 | 0.049 | 0.19 | 0.04 | 0.99 |
|  | Physical activity | 2-3 times/week | -1.58 | 0.75 | 4.47 | 0.034 | 0.20 | 0.05 | 0.89 |
|  |  | ≥ 4 times/week | -1.71 | 0.94 | 3.32 | 0.068 | 0.18 | 0.03 | 1.13 |
|  | Education level | High school | 1.83 | 0.73 | 6.22 | 0.013 | 6.26 | 1.48 | 26.01 |
| 10-13 |  |  |  |  |  |  |  |  |  |
|  | Child NW |  | 1.10 | 0.30 | 13.30 | 0.000 | 3.07 | 1.68 | 5.61 |
|  | Nap | Sometimes | -0.89 | 0.44 | 4.02 | 0.045 | 0.40 | 0.17 | 0.90 |
|  | Physical activity | 2-3 times/week | -0.91 | 0.50 | 3.33 | 0.064 | 0.40 | 0.14 | 1.06 |
|  | Mother’s age |  | -0.97 | 0.04 | 5.38 | 0.020 | 0.90 | 0.83 | 0.98 |
|  | Education level | High school | 1.18 | 0.54 | 4.77 | 0.028 | 3.28 | 1.13 | 9.49 |
| 6 - 9 |  |  |  |  |  |  |  |  |  |
|  | Child NW |  | 0.91 | 0.31 | 8.57 | 0.003 | 2.48 | 1.35 | 4.58 |
|  | Physical activity | 2-3 times/week | -1.05 | 0.50 | 4.29 | 0.038 | 0.35 | 0.13 | 0.94 |
|  |  | ≥ 4 times/week | -1.37 | 0.66 | 4.25 | 0.039 | 0.25 | 0.07 | 0.92 |
|  | Education level | High school | 1.29 | 0.55 | 5.47 | 0.019 | 3.64 | 1.23 | 10.70 |

Finally, as for the other groups of mothers, the childreńs awakenings continue to impact on the motherś sleep quality in all categories, where a higher number decreased the probability of being in the category of PSQI_0-5_.

## Discussion

In the present report we evaluated maternal sleep quality in women from delivery until the first 10 years of life of their child. In addition, we analyzed possible predictors that may account for sleep quality at different stages. We found that maternal sleep quality was poor for all mothers regardless of the age of the child. Particularly, it was especially low during the first two years, with a significant improvement towards age 3, returning to lower levels at ages 4 and 5 of the child, to finally improve towards the ages of 8 to 10 of the child. For most mothers, personal factors such as the level of education and performing physical activity impact on their sleep quality. Regarding the sleep characteristics of the child, the number of awakenings affected maternal sleep quality at all stages of the child. However, for the mothers with infant-toddlers and preschool children, additional aspects of their children’s sleep, as well as social and nocturnal family dynamic, also predict their sleep quality.

### Maternal sleep quality during the first 12 months after childbirth

We found that the average PSQI of mothers is particularly high across all ages of the child studied. Specifically, during the first 12 months after delivery, we have found that the median PSQI of the mothers is 10, with 88 % of them reporting poor sleep quality (PSQI > 5). Reports about sleep quality using the PSQI during the postpartum period differ according to the population, but they all agree that most mothers have poor sleep quality. Similarly to our data, an article from Australia of women with babies from 0 to 12 months showed and average PSQI of 9.63 [44]. Also, a report from a population of Canada, described four groups of maternal sleep according to their sleep quality with two of these groups with an average PSQI of 9-11 at 3 and 6 months after delivery [45]. European articles have quite different PSQI values: a population of Spanish mothers during their first 6 months of lactation had a median much higher compared to ours results, finding a PSQI of 19 [40], while a report from a population of Italy and Switzerland shows that poor quality of sleep was reported by 71% of women in the first month after delivery, but with PSQI values much lower than ours, about 5-8 from birth to 6 months [36]. An Asian article from Iran shows a PSQI varying 7.5-8.5 in the first two months after childbirth [46], similar to one in United States of America (7.3-7.7) [47]. The only report we found from a South American country is from Brazil and showed that approximately 70% of postpartum women from 1 to 73 days after delivery had poor sleep quality (PSQI > 5), but they did not show the average value of the PSQI [34]. Thus, our data is in accordance with many articles, but PSQI values during postpartum are still highly variable across different countries.

### Maternal sleep quality beyond first year after childbirth

Prior studies have focused on sleep disturbances during the postpartum period. However, little is known about the sleep quality of mothers beyond the postpartum period. Here we show how sleep quality varies during the first 10 years of motherhood. Specifically, after the postpartum period the PSQI continued to be elevated during the first two years of life of the child, beginning to improve at the stage of three years. However, PSQI values of mothers with children of 4-5 and 6-7 years old did not differ significantly compared to the group of mothers with children from 0 to 5 months old, suggesting a return to a poorer sleep quality that seems to recover in the group of mothers with children of 8-10 years old. These data are in accordance with an article about a population of German mothers, which shows that maternal sleep satisfaction and duration did not fully recovery for up to 6 years after childbirth [15]. Another German report shows that mothers with children from 0 to 4 years with sleep problems have a mean value of PSQI around 8, but PSQI range is 4-6 in mothers that report their child not having sleep problems [48]. Although these values are much lower than ours, Uruguayan population already experience high scores in the PSQI [43]. Another large-scale study showed that maternal sleep quality from 13 different countries described that 55% of mothers with children from 0-6 years (average of 2 years) reported poor sleep. Although they used the PSQI, the average value is not described or differentiated according to the age [37]. In regard to this data, our findings show that many more mothers experience poor sleep (around 80-85%), in agreement with the fact that Uruguayan population has poorer sleep quality compared to many other countries [32].

### Protective and risk factors on maternal sleep quality

Most sleep research has focused mainly on high-income nations [32, 33]. However, several sociocultural factors impact in aspects of sleep, such as sleep onset and sleep duration [32]. It might be expected that certain variables that contribute to poor sleep quality could also vary from one population to another.

One of the predictors at all stages of motherhood for bad quality of sleep was child’s nocturnal awakenings. In concordance with our results, a German report that analyzed mothers with children up to 4 years shows that maternal sleep quality depends on the child’s sleep quality [48]. In addition, in mothers of children from 0 to 6 years, Mindell and col. (2015) reported that the child’s sleep pattern significantly impacts maternal sleep, finding a positive correlation between child’s awakenings and motheŕs awakenings, that was stronger in younger children (0 to 36 months) than in older ones (37-72 months) [37]. However, we found that the odds of having poor sleep quality according to the child’s awakenings were higher in mothers with school children (2.4-3 more probability) than in mothers with infant-toddlers (1.3-1.8 more probability). In addition, our current data demonstrates for the first time that nighttime awakenings in children up to 10 years of age also affect maternal sleep.

Another variable that increased the probability of poorer maternal sleep quality at all ages of the child was lower levels of education compared to complete college education. It has already been described that sleep quality varies according to the level of education, being better in more highly educated individuals, not only in general population [49], but also in postpartum women [41].

Regarding the social environment, several reports show that co-sleeping negatively impacts on the motheŕs sleep quality during the postpartum period [16, 18, 50]. The present data reveal that co-sleeping not only affects mothers with infants and toddlers but also with preschool children. Specifically, using polysomnography, Mosko et al. (1997) evidence that mothers co-sleeping with their 2-3-month babies slightly reduces stage 3 and 4 of NREM sleep and have more frequent arousals, indicating a poorer sleeping compared to solitary sleepers [18]. In addition, Stremler et al. (2013) using actigraphy show similar results [50]. Our current data is in line with these previous data, where we found that co-sleeping arrangement increases the probability for mothers of having a bad sleep quality compared to solitary sleepers. From the evolutionary point of view, this reduction in maternal sleep quality in co-sleeping arrangement should be seen as beneficial for infants. For instance, more frequent arousals would increase mothers’ opportunities to monitor their child [18]. In this sense, co-sleeping may have a differential impact on maternal sleep quality depending on whether the child has achieved “sleeping through the night” or experiences sleep disturbances. Hence, we hypothesized that mothers whose child has sleep disturbances, co-sleeping might improve maternal sleep quality, as the duration of the arousal episodes may be diminished. However, new studies must be carried out to prove this hypothesis.

We found that the collaboration of another adult during child’s nocturnal awakenings predicts worse sleep quality in the group of mothers with infant-toddler children with higher PSQI (PSQI_14-21_), but not in the other groups [16, 18]. We believe that the collaboration factor may indirectly influence maternal sleep quality. Specifically, we hypothesize that mothers who experience worse sleep quality due to childreńs nighttime awakenings may be more likely to seek support during these episodes, which could explain this seemingly contradictory association. In this regard, it has been discussed that maternal sleep pattern may be shaped by the nocturnal needs of the child, as these are usually managed by the mother [18], finding stronger associations in mother-child than father-child in various sleep patterns [51]. In families with older children, the proportion of children who do not require nocturnal care progressively increased (from 7% in infant toddlers, 19.7 % in preschool to 44.8 % in school children). These changes may account for the lack of association between nocturnal collaboration and maternal sleep quality in families with older children.

Maternal age differentially impacted sleep quality in the group of mothers, where old age in the preschool group was a risk factor while in school group it was a protective factor. The fact that mothers of preschool children were significantly younger than the ones with school children may account for this difference. In this sense, the range of age of older mothers in the group with preschool children overlaps with those of younger mothers in the group with school children. Thus, that range of age is what increases the probability of having bad sleep quality. In this regard, it has been reported that too young (under 18) or too old (above 35) mothers during the postpartum period (three months after delivery) were associate with sleep disturbances [34]. However, it is important to highlight that the age of the mother in our results was significant only in one of the three PSQI categories of bad quality of sleep in each group of mothers. Accordingly, it has been reported that parental age was unrelated or weakly related to sleep quality [15].

### Limitations

Firstly, we assess sleep quality subjectively. Future studies using actigraphy are needed to provide more accurate data. However, the current methodology allowed us to include more participants from different areas of the country (approximately half were from the capital and the rest from other areas of the country). Also, other variables not analyzed in this report may have an impact on maternal sleep quality.

Furthermore, although most previous research has considered the effects of breastfeeding on maternal sleep [11, 15, 52], with inconsistent findings, we could not analyzed the relationship between infant feeding methods and sleep quality, as the number of mothers who only bottle-fed were only 4, compared to 146 mothers who breastfed their infants from 0 to 5 months.

Lastly, the proportion of participants with complete college education in our sample was high compared to the entire population of the country; hence, it is not representative of our population. This factor may have contributed to lower PSQI scores than would have been observed if mothers from other communities in our country had completed the questionnaire [53]. It is worth noting that other studies on maternal sleep have analyzed populations with similar educational levels [37, 45].

### Conclusion and future research

The current findings provide empirical support to the popular knowledge that maternal sleep quality disturbances extend well beyond the postpartum period. Given the diverse cultural factors that can improve or worsen the quality of sleep, it is essential to conduct future research about maternal sleep beyond the postpartum period in diverse cultural contexts. Additionally, raising awareness among general public and health professionals is crucial to better support women experiencing sleep disturbances throughout motherhood. As far as we know, this is the first report that focuses on maternal sleep during the first 10 years of motherhood.

## Data Availability

All data produced in the present work are contained in the manuscript

## References

1. Hunter, L.P., J.D. Rychnovsky, and S.M. Yount, A selective review of maternal sleep characteristics in the postpartum period. J Obstet Gynecol Neonatal Nurs, 2009. 38(1): p. 60–8.

2. Lyamin, O., et al., Behavioral aspects of sleep in bottlenose dolphin mothers and their calves. Physiol Behav, 2007. 92(4): p. 725–33.

3. Sivadas, N., et al., Dynamic changes in sleep pattern during post-partum in normal pregnancy in rat model. Behav Brain Res, 2016. 320: p. 264–274.

4. Benedetto, L., et al., A descriptive analysis of sleep and wakefulness states during maternal behaviors in postpartum rats. Arch Ital Biol, 2017. 155(3): p. 99–109.

5. Stremler, R., K. Sharkey, and A. Wolfson, *Postpartum Period and Early Motherhood*, in Principles and Practice of Sleep Medicine, M.H. Kryger, T. Roth, and W.C. Dement, Editors. 2017, Elsevier: Philadelphia, PA. p. 1547–1552.

6. Lee, K.A., Alterations in sleep during pregnancy and postpartum: a review of 30 years of research. Sleep Med Rev, 1998. 2(4): p. 231–42.

7. Blyton, D.M., C.E. Sullivan, and N. Edwards, Lactation is associated with an increase in slow-wave sleep in women. J Sleep Res, 2002. 11(4): p. 297–303.

8. Thomas, K.A. and R.L. Burr, Melatonin level and pattern in postpartum versus nonpregnant nulliparous women. J Obstet Gynecol Neonatal Nurs, 2006. 35(5): p. 608–15.

9. Montgomery-Downs, H.E., et al., Normative longitudinal maternal sleep: the first 4 postpartum months. Am J Obstet Gynecol, 2010. 203(5): p. 465 e1-7.

10. Nishihara, K., et al., Delta and theta power spectra of night sleep EEG are higher in breast-feeding mothers than in non-pregnant women. Neurosci Lett, 2004. 368(2): p. 216–20.

11. Gay, C.L., K.A. Lee, and S.Y. Lee, Sleep patterns and fatigue in new mothers and fathers. Biol Res Nurs, 2004. 5(4): p. 311–8.

12. Nishihara, K. and S. Horiuchi, Changes in sleep patterns of young women from late pregnancy to postpartum: relationships to their infants’ movements. Percept Mot Skills, 1998. 87(3 Pt 1): p. 1043-56.

13. Signal, T.L., et al., Sleep duration and quality in healthy nulliparous and multiparous women across pregnancy and post-partum. Aust N Z J Obstet Gynaecol, 2007. 47(1): p. 16–22.

14. Wu, J., et al., Association between sleep quality and physical activity in postpartum women. Sleep Health, 2019. 5(6): p. 598–605.

15. Richter, D., et al., Long-term effects of pregnancy and childbirth on sleep satisfaction and duration of first-time and experienced mothers and fathers. Sleep, 2019. 42(4).

16. Volkovich, E., et al., Mother-infant sleep patterns and parental functioning of room-sharing and solitary-sleeping families: a longitudinal study from 3 to 18 months. Sleep, 2018. 41(2).

17. Stremler, R. and A. Wolfson, The postpartum period, in Principles and Practice of Sleep Medicine, M.H. Kryger, T. Roth, and W.C. Dement, Editors. 2011, Elsevier: Canada. p. 1587–1591.

18. Mosko, S., C. Richard, and J. McKenna, Maternal sleep and arousals during bedsharing with infants. Sleep, 1997. 20(2): p. 142–50.

19. Pennestri, M.H., et al., Uninterrupted Infant Sleep, Development, and Maternal Mood. Pediatrics, 2018. 142(6).

20. Kalogeropoulos, C., et al., Sleep patterns and intraindividual sleep variability in mothers and fathers at 6 months postpartum: a population-based, cross-sectional study. BMJ Open, 2022. 12(8): p. e060558.

21. Henderson, J.M., K.G. France, and N.M. Blampied, The consolidation of infants’ nocturnal sleep across the first year of life. Sleep Med Rev, 2011. 15(4): p. 211–20.

22. Field, T., Infant sleep problems and interventions: A review. Infant Behav Dev, 2017. 47: p. 40–53.

23. Kuhn, B.R. and A.J. Elliott, Treatment efficacy in behavioral pediatric sleep medicine. J Psychosom Res, 2003. 54(6): p. 587–97.

24. Archbold, K.H., et al., Symptoms of sleep disturbances among children at two general pediatric clinics. J Pediatr, 2002. 140(1): p. 97–102.

25. Owens, J.A., et al., Sleep habits and sleep disturbance in elementary school-aged children. J Dev Behav Pediatr, 2000. 21(1): p. 27–36.

26. Richman, N., J.E. Stevenson, and P.J. Graham, Prevalence of behaviour problems in 3-year-old children: an epidemiological study in a London borough. J Child Psychol Psychiatry, 1975. 16(4): p. 277–87.

27. Richman, N., A community survey of characteristics of one- to two-year-olds with sleep disruptions. J Am Acad Child Psychiatry, 1981. 20(2): p. 281–91.

28. Ramchandani, P., et al., A systematic review of treatments for settling problems and night waking in young children. BMJ, 2000. 320(7229): p. 209-13.

29. Bax, M.C., Sleep disturbance in the young child. Br Med J, 1980. 280(6224): p. 1177-9.

30. Anders, T.F. and L.A. Eiben, Pediatric sleep disorders: a review of the past 10 years. J Am Acad Child Adolesc Psychiatry, 1997. 36(1): p. 9–20.

31. Boergers, J., et al., Child sleep disorders: associations with parental sleep duration and daytime sleepiness. J Fam Psychol, 2007. 21(1): p. 88–94.

32. Coutrot, A., et al., Reported sleep duration reveals segmentation of the adult life-course into three phases. Nature Communications, 2022. 13(1): p. 7697.

33. Simonelli, G., et al., Sleep health epidemiology in low and middle-income countries: a systematic review and meta-analysis of the prevalence of poor sleep quality and sleep duration. 2018(2352-7226 (Electronic)).

34. Motta, A.J.P., et al., Factors Associated with Poor Sleep Quality in Postpartum Women: A Crossectional Study. Sleep Sci, 2024. 17(3): p. e263–e271.

35. Tikotzky, L., et al., A longitudinal study of the links between maternal and infant nocturnal wakefulness. Sleep Health, 2022. 8(1): p. 31–38.

36. Manconi, M., et al., Sleep and sleep disorders during pregnancy and postpartum: The Life-ON study. Sleep Med, 2024. 113: p. 41–48.

37. Mindell, J.A., et al., Relationship Between Child and Maternal Sleep: A Developmental and Cross-Cultural Comparison. J Pediatr Psychol, 2015. 40(7): p. 689–96.

38. Buysse, D.J., et al., The Pittsburgh Sleep Quality Index: a new instrument for psychiatric practice and research. Psychiatry Res, 1989. 28(2): p. 193–213.

39. Hita-Contreras, F., et al., Reliability and validity of the Spanish version of the Pittsburgh Sleep Quality Index (PSQI) in patients with fibromyalgia. Rheumatol Int, 2014. 34(7): p. 929–36.

40. Cabrera-Dominguez, G., et al., Women during Lactation Reduce Their Physical Activity and Sleep Duration Compared to Pregnancy. Int J Environ Res Public Health, 2022. 19(18).

41. Gessesse, D.N., et al., Prevalence and associated factors of poor sleep quality among postpartum women in North West Ethiopia: a community-based study. BMC Psychiatry, 2022. 22(1): p. 538.

42. Gao, M., et al., Association of sleep quality during pregnancy with stress and depression: a prospective birth cohort study in China. BMC Pregnancy Childbirth, 2019. 19(1): p. 444.

43. Torterolo, P., et al., Calidad de sueño en Uruguay al inicio de la pandemia Sleep quality in Uruguay at the beginning of the pandemic Anales de la Facultad de Medicina, 2023. 10(2).

44. Newman, L., et al., How do infant feeding method, sleeping location, and postpartum depression interact with maternal sleep quality? Sleep Med, 2023. 110: p. 183–189.

45. Tomfohr, L.M., et al., Trajectories of Sleep Quality and Associations with Mood during the Perinatal Period. Sleep, 2015. 38(8): p. 1237–45.

46. Keshavarz Afshar, M., et al., Lavender fragrance essential oil and the quality of sleep in postpartum women. Iran Red Crescent Med J, 2015. 17(4): p. e25880.

47. Christian, L.M., et al., Sleep quality across pregnancy and postpartum: effects of parity and race. Sleep Health, 2019. 5(4): p. 327–334.

48. Lollies, F., et al., Child Sleep Problems Affect Mothers and Fathers Differently: How Infant and Young Child Sleep Affects Paternal and Maternal Sleep Quality, Emotion Regulation, and Sleep-Related Cognitions. Nat Sci Sleep, 2022. 14: p. 137–152.

49. Grandner, M.A., et al., Sleep disparity, race/ethnicity, and socioeconomic position. Sleep Med, 2016. 18: p. 7–18.

50. Stremler, R., et al., Infant Sleep Location: Bed Sharing, Room Sharing and Solitary Sleeping at 6 and 12 Weeks Postpartum. The Open Sleep Journal, 2013. **6**(M10): p. 77-86.

51. Zhang, J., et al., Roles of parental sleep/wake patterns, socioeconomic status, and daytime activities in the sleep/wake patterns of children. J Pediatr, 2010. 156(4): p. 606–12 e5.

52. Doan, T., et al., Breast-feeding increases sleep duration of new parents. J Perinat Neonatal Nurs, 2007. 21(3): p. 200–6.

53. McEvoy, K.M., et al., Poor Postpartum Sleep Quality Predicts Subsequent Postpartum Depressive Symptoms in a High-Risk Sample. J Clin Sleep Med, 2019. 15(9): p. 1303–1310.

