## Supplemental Tables for "Maternal sleep during the first ten years of the child’s life": Table Supp.pdf

**Table S1.** Variables included in the analysis of sleep quality of mothers with infants-toddlers children (0–2 years).

| PSQI category | Variable | Variable sub-category | B | Standard Error | Wald | *P* | OR | CI 95% for OR | |
|---|---|---|---|---|---|---|---|---|---|
| | | | | | | | | *Low Bound* | *Upper Bound* |
| **14 – 21** | | | | | | | | | |
| | Mother´s age | | 0.04 | 0.04 | 1.10 | 0.294 | 1.03 | 0.96 | 1.11 |
| | **Education level** | School | 1.30 | 0.89 | 2.12 | 0.155 | 3.68 | 0.63 | 21.30 |
| | | **High school** | 0.77 | 0.40 | 3.69 | **0.055** | 2.17 | 0.98 | 4.79 |
| | Work | Unemployed | 0.49 | 0.50 | 0.96 | 0.326 | 1.64 | 0.61 | 4.42 |
| | **Family composition** | Extended | -0.09 | 0.83 | 0.01 | 0.988 | 0.98 | 0.17 | 4.67 |
| | | **Single** | 1.67 | 0.81 | 4.25 | **0.039** | 5.34 | 1.08 | 26.21 |
| | Nº children | | -0.28 | 0.24 | 1.35 | 0.244 | 0.75 | 0.46 | 1.21 |
| | Child Sex | Female | 0.20 | 0.31 | 0.42 | 0.516 | 1.23 | 0.65 | 2.23 |
| | **Night collaboration** | **No require** | -1.94 | 1.10 | 3.12 | **0.077** | 0.14 | 0.16 | 1.23 |
| | | **Sometimes** | 1.34 | 0.45 | 8.64 | **0.003** | 3.84 | 1.56 | 9.43 |
| | | Always | 0.49 | 0.39 | 1.57 | 0.209 | 1.63 | 0.76 | 3.53 |
| | **Child NW** | | 0.68 | 0.12 | 23.10 | **0.000** | 1.83 | 1.38 | 2.07 |
| | Sleep arrangements | SS | -0.63 | 0.45 | 1.95 | 0.162 | 0.53 | 0.22 | 1.28 |
| | | RS | -0.56 | 0.38 | 2.18 | 0.140 | 0.57 | 0.27 | 1.20 |
| | **Nap** | **Always** | -1.36 | 0.66 | 4.12 | **0.041** | 0.26 | 0.07 | 0.95 |
| | | Frequently | -0.11 | 0.65 | 0.03 | 0.856 | 0.89 | 0.25 | 3.15 |
| | | Sometimes | -0.36 | 0.35 | 1.02 | 0.311 | 0.69 | 0.35 | 1.39 |
| | **Physical activity** | ≥ 4 times/week | -0.20 | 0.82 | 0.06 | 0.805 | 0.81 | 0.16 | 4.11 |
| | | 2 - 3 times/week | -1.35 | 0.43 | 9.57 | **0.002** | 0.25 | 0.11 | 0.61 |
| | | 1 time/week | -0.66 | 0.45 | 0.02 | 0.886 | 0.93 | 0.38 | 2.23 |
| **10- 13** | | | | | | | | | |
| | Mother´s age | | -0.40 | 0.29 | 1.83 | 0.176 | 0.96 | 0.90 | 1.01 |
| | Education level | School | 0.77 | 0.75 | 1.05 | 0.304 | 2.17 | 0.49 | 9.50 |
| | | High school | 0.18 | 0.34 | 0.28 | 0.593 | 1.19 | 0.61 | 2.32 |
| | Work | Unemployed | -0.34 | 0.44 | 0.59 | 0.440 | 0.71 | 0.29 | 1.70 |
| | Family composition | Extended | -0.57 | 0.70 | 0.69 | 0.414 | 0.56 | 0.14 | 2.23 |
| | | Single | 1.02 | 0.72 | 1.97 | 0.160 | 2.77 | 0.66 | 11.50 |

| | | | | | | | | | |
|---|---|---|---|---|---|---|---|---|---|
| | Nº children | | -0.76 | 0.19 | 0.15 | 0.693 | 0.92 | 0.63 | 1.35 |
| | Child Sex | Female | 0.29 | 0.25 | 1.35 | 0.244 | 1.34 | 0.81 | 2.21 |
| | Night collaboration | No require | -0.73 | 0.47 | 2.42 | 0.120 | 0.47 | 0.19 | 1.21 |
| | | Sometimes | 0.37 | 0.39 | 0.89 | 0.344 | 1.44 | 0.67 | 3.11 |
| | | Always | 0.15 | 0.31 | 0.22 | 0.634 | 1.16 | 0.63 | 2.13 |
| | **Child NW** | | 0.53 | 0.10 | 26.10 | **0.000** | 1.69 | 1.37 | 2.06 |
| | **Sleep arrangements** | **SS** | -0.77 | 0.36 | 4.64 | **0.031** | 0.46 | 0.30 | 0.93 |
| | | RS | -0.22 | 0.30 | 0.52 | 0.471 | 0.80 | 0.43 | 1.46 |
| | Nap | Always | 0.11 | 0.42 | 0.07 | 0.787 | 1.12 | 0.49 | 2.56 |
| | | Frequently | -0.32 | 0.54 | 0.37 | 0.546 | 0.72 | 0.24 | 2.09 |
| | | Sometimes | 0.00 | 0.28 | 0.00 | 0.988 | 1.00 | 0.57 | 1.75 |
| | **Physical activity** | ≥ 4 times/week | 0.87 | 0.60 | 0.02 | 0.886 | 1.09 | 0.33 | 3.55 |
| | | **2-3 times/week** | -0.60 | 0.30 | 4.0 | **0.045** | 0.55 | 0.30 | 0.98 |
| | | 1 time/week | 0.21 | 0.38 | 0.32 | 0.572 | 1.23 | 0.58 | 2.60 |
| **6 - 9** | | | | | | | | | |
| | Mother´s age | | 0.02 | 0.02 | 0.72 | 0.405 | 1.02 | 0.96 | 1.08 |
| | Education level | School | -0.43 | 0.90 | 0.23 | 0.628 | 0.64 | 0.11 | 3.78 |
| | | High school | 0.25 | 0.33 | 0.57 | 0.448 | 1.28 | 0.66 | 2.48 |
| | Work | Unemployed | -0.55 | 0.46 | 1.42 | 0.232 | 0.57 | 0.23 | 1.42 |
| | Family composition | Extended | 0.09 | 0.63 | 0.02 | 0.877 | 1.10 | 0.31 | 3.83 |
| | | Single | 0.86 | 0.72 | 1.43 | 0.231 | 2.37 | 0.57 | 9.74 |
| | Nº children | | -0.29 | 0.19 | 2.33 | 0.126 | 0.74 | 0.51 | 1.08 |
| | Child Sex | Female | 0.33 | 0.25 | 1.80 | 0.179 | 1.40 | 0.86 | 2.29 |
| | **Night collaboration** | **No require** | -0.83 | 0.46 | 3.33 | **0.067** | 0.43 | 0.17 | 1.06 |
| | | Sometimes | 0.48 | 0.38 | 1.58 | 0.208 | 1.62 | 0.76 | 3.44 |
| | | Always | 0.22 | 0.30 | 0.51 | 0.474 | 1.24 | 0.68 | 2.28 |
| | **Child NW** | | 0.33 | 0.10 | 10.20 | **0.001** | 1.39 | 1.13 | 1.69 |
| | Sleep arrangements | SS | -0.51 | 0.34 | 2.22 | 0.136 | 0.59 | 0.30 | 1.17 |
| | | RS | -0.26 | 0.30 | 0.70 | 0.401 | 0.77 | 0.42 | 1.41 |
| | Nap | Always | 0.88 | 0.42 | 0.43 | 0.836 | 1.09 | 0.47 | 2.50 |
| | | Frequently | -0.40 | 0.54 | 0.55 | 0.459 | 0.66 | 0.23 | 1.93 |
| | | Sometimes | -0.00 | 0.28 | 0.00 | 0.984 | 0.99 | 0.57 | 1.72 |

| | | | | | | | | | |
|---|---|---|---|---|---|---|---|---|---|
| | Physical activity | ≥ 4 times/week | -0.38 | 0.64 | 0.37 | 0.541 | 0.67 | 0.19 | 2.35 |
| | | 2-3 times/week | -0.40 | 0.29 | 1.91 | 0.167 | 1.67 | 0.37 | 1.18 |
| | | 1 time/ week | 0.51 | 0.37 | 1.92 | 0.166 | 1.67 | 0.80 | 3.45 |

Multinomial logistic regression in the group of mothers with infant-toddlers children, showing all variables analyzed. The reference category for the dependent variable was PSQI < 5. Variables that had a statistical tendency or were significant are written in bold letters. Reference categories for the independent variables were as follows: for education level, *complete college*; for work, employed; for family composition, *nuclear family*; for child sex, male; for night collaboration, *does not have*; for sleep arrangements, *bed-sharing*; for napping, *never taking naps*; for physical activity (≥ 30 min), *never does*. **Model fit:** $R^2$ = 0.21 (Cox & Snell), 0.23 (Nagelkerke); Model $\chi^2$ = 173.61, $p < 0.05$. Abbreviations**:** B, beta coefficient; CI, confidence interval; NW, night awakening; OR, odds ratio; RS, room sharing; SS, solitary sleep.

**Table S2. Variables included in the analysis of sleep quality in mothers** with preschool children (3-5 years).

| **PSQI category** | **Variable** | **Variable sub-category** | **B** | **Standard Error** | **Wald** | ***P*** | **OR** | **CI 95% for OR** | |
|---|---|---|---|---|---|---|---|---|---|
| | | | | | | | | ***Low Bound*** | ***Upper Bound*** |
| **14 - 21** | | | | | | | | | |
| | Mother´s age | | 0.10 | 0.07 | 2.23 | 0.138 | 1.11 | 0.96 | 1.27 |
| | Education level | School* | -16.2 | 6164.67 | 0.00 | 0.998 | 9.09E-008 | 0.00 | - |
| | | High school | 1.27 | 0.84 | 2.25 | 0.133 | 3.57 | 0.67 | 18.8 |
| | Work | Unemployed | -0.44 | 0.99 | 0.19 | 0.659 | 0.64 | 0.09 | 4.56 |
| | Family composition | Extended* | -20.0 | 6936.2 | 0.00 | 0.998 | 1.87E-009 | 0.00 | - |
| | | Single | -0.57 | 0.93 | 0.38 | 0.536 | 0.56 | 0.09 | 3.94 |
| | Nº children | | -0.24 | 0.46 | 0.00 | 0.959 | 0.97 | 0.39 | 2.42 |
| | Child sex | Female | 0.16 | 0.64 | 0.06 | 0.797 | 1.17 | 0.33 | 4.11 |
| | Night collaboration | No require | 1.09 | 0.79 | 1.87 | 0.171 | 2.97 | 0.63 | 14.1 |
| | | Sometimes | 0.23 | 0.91 | 0.06 | 0.796 | 1.26 | 0.21 | 7.62 |
| | | Always | 0.01 | 0.82 | 0.00 | 0.985 | 1.01 | 0.20 | 5.15 |
| | **Child NW** | | 1.12 | 0.34 | 12.30 | **0.000** | 3.32 | 1.72 | 6.44 |
| | **Sleep arrangements** | **SS** | -1.48 | 0.71 | 4.30 | **0.038** | 0.22 | 0.56 | 0.92 |
| | | RS | -1.78 | 1.32 | 1.83 | 0.176 | 0.16 | 0.13 | 2.22 |
| | Nap | Always* | -16.89 | 5697.2 | 0.00 | 0.998 | 4.60E-008 | 0.00 | - |
| | | Frequently | 0.15 | .00 | - | - | 1.16 | 1.16 | 1.17 |
| | | Sometimes | 0.61 | 0.69 | 0.80 | 0.370 | 1.85 | 0.48 | 7.17 |

| | | | | | | | | | |
|---|---|---|---|---|---|---|---|---|---|
| | **Physical activity** | ≥ 4 times/week* | -18.56 | 6424.7 | 0.00 | 0.998 | 8.66E-009 | 0.00 | - |
| | | **2 - 3 times/week** | -1.50 | 0.85 | 3.19 | **0.074** | 0.22 | 0.042 | 1.15 |
| | | 1 time/week | 1.28 | 0.78 | 2.67 | 0.102 | 3.61 | 0.77 | 16.8 |
| **10- 13** | | | | | | | | | |
| | Mother´s age | | 0.06 | 0.50 | 2.22 | 0.146 | 1.07 | 0.97 | 1.17 |
| | Education level | School | 1.72 | 1.27 | 1.83 | 0.176 | 5.61 | 0.46 | 68.0 |
| | | High school | 0.59 | 0.56 | 1.10 | 0.233 | 2.97 | 0.60 | 5.44 |
| | **Work** | Unemployed | -1.31 | 0.73 | 3.21 | **0.073** | 0.26 | 0.091 | 4.56 |
| | Family composition | Extended | -0.57 | 1.06 | 0.29 | 0.588 | 0.56 | 0.00 | - |
| | | Single | 0.11 | 0.67 | 0.03 | 0.860 | 1.12 | 0.09 | 3.49 |
| | Nº children | | 0.34 | 0.31 | 1.22 | 0.269 | 1.41 | 0.39 | 2.42 |
| | Child Sex | Female | -0.36 | 0.40 | 0.00 | 0.938 | 0.96 | 0.60 | 3.06 |
| | Night collaboration | No require | -0.14 | 0.59 | 0.05 | 0.809 | 0.86 | 0.62 | 14.1 |
| | | Sometimes | 0.39 | 0.57 | 0.47 | 0.492 | 1.48 | 0.21 | 7.62 |
| | | Always | 0.24 | 0.51 | 0.21 | 0.644 | 1.27 | 0.20 | 5.15 |
| | **Child NW** | | 0.55 | 0.22 | 5.83 | **0.016** | 1.73 | 1.11 | 2.71 |
| | **Sleep arrangements** | **SS** | -1.48 | 0.52 | 7.76 | **0.005** | 0.23 | 0.08 | 0.66 |
| | | **RS** | -2.07 | 0.78 | 6.96 | **0.008** | 0.12 | 0.03 | 0.57 |
| | Nap | Always | 1.09 | 0.91 | 1.42 | 0.232 | 2.97 | 0.49 | 17.73 |
| | | Frequently* | 18.3 | 5408.4 | 0.00 | 0.997 | 95147866.03 | 0.00 | - |
| | | Sometimes | 0.60 | 0.49 | 1.51 | 0.219 | 1.83 | 0.69 | 4.83 |
| | **Physical activity** | ≥ 4 times/week | -0.81 | 0.85 | 0.91 | 0.339 | 0.44 | 0.08 | 2.34 |
| | | **2-3 times/week** | -1.13 | 0.48 | 5.54 | **0.019** | 0.32 | 0.12 | 0.82 |
| | | 1 time/week | -0.84 | 0.59 | 1.97 | 0.160 | 0.43 | 0.13 | 1.39 |
| **6 – 9** | | | | | | | | | |
| | **Mother´s age** | | 0.13 | 0.04 | 7.91 | **0.005** | 1.14 | 0.80 | 2.37 |
| | **Education level** | **School** | 2.16 | 1.27 | 2.86 | **0.091** | 8.69 | 0.70 | 106.72 |
| | | High school | 0.85 | 0.57 | 2.22 | 0.136 | 2.36 | 0.76 | 7.23 |
| | Work | Unemployed | -0.49 | 0.69 | 0.52 | 0.472 | 0.60 | 0.16 | 2.35 |
| | Family composition | Extended | -0.99 | 1.11 | 0.78 | 0.374 | 0.37 | 0.04 | 3.31 |
| | | Single | -0.77 | 0.70 | 0.01 | 0.913 | 0.92 | 0.23 | 3.66 |

| | | | | | | | | | |
|---|---|---|---|---|---|---|---|---|---|
| | Nº children | | 0.39 | 0.31 | 1.63 | 0.201 | 1.48 | 0.80 | 2.74 |
| | Child sex | Female | 0.30 | 0.41 | 0.53 | 0.465 | 1.35 | 0.60 | 3.06 |
| | Night collaboration | No require | 0.41 | 0.58 | 0.49 | 0.483 | 1.50 | 0.48 | 4.76 |
| | | Sometimes | 0.88 | 0.61 | 0.20 | 0.885 | 1.09 | 0.33 | 3.63 |
| | | Always | 0.22 | 0.53 | 0.18 | 0.666 | 1.25 | 0.44 | 3.55 |
| | Child NW | | 0.27 | 0.24 | 1.33 | 0.249 | 1.31 | 0.83 | 2.10 |
| | Sleep arrangements | SS | -0.42 | 0.56 | 0.54 | 0.461 | 0.65 | 0.21 | 2.00 |
| | | RS | -0.73 | 0.79 | 0.85 | 0.356 | 0.48 | 0.10 | 2.28 |
| | Nap | Always | 1.00 | 0.95 | 1.11 | 0.291 | 2.74 | 0.42 | 17.74 |
| | | Frequently* | 19.1 | 5408.4 | 0.00 | 0.997 | 204491050.52 | 0.00 | - |
| | | Sometimes | 0.72 | 0.50 | 2.06 | 0.150 | 2.07 | 0.77 | 5.60 |
| | **Physical activity** | ≥ 4 times/week | -1.39 | 0.97 | 2.03 | 0.154 | 0.25 | 0.03 | 1.68 |
| | | **2-3 times/week** | -0.89 | 0.49 | 3.31 | **0.069** | 0.41 | 0.16 | 1.07 |
| | | 1 time/week | -0.80 | 0.62 | 1.68 | 0.194 | 0.45 | 0.13 | 1.50 |

Multinomial logistic regression in the group of mothers with preschool children, showing all variables analyzed. The reference category for the dependent variable was PSQI <5. Variables that had a statistical tendency or were significant are written in bold letters. Reference categories for the independent variables were as follows: for education level, *complete college*; for work, employed; for family composition, *nuclear family*; for child sex, male; for night collaboration, *does not have*; for sleep arrangements, *bed-sharing*; for napping, *never taking naps*; for physical activity (≥ 30 min), *never does.* Subcategories marked with an asterisk (*), the number of mothers was too low for statistical interpretation. Model fit: $R^2$ = 0.31 (Cox & Snell), 0.34 (Nagelkerke); Model $\chi^2$ = 90.61, $p$ < 0.05. Abbreviations: B, beta coefficient; CI, confidence interval; NW, night awakening; OR, odds ratio; RS, room sharing; SS, solitary sleep.

**Table S3. Variables included in the analysis of sleep quality in mothers** with school children (6-10 years).

| **PSQI category** | **Variable** | **Variable sub-category** | **B** | **Standard Error** | **Wald** | ***P*** | **OR** | **CI 95% for OR** *Low Bound* | *Upper Bound* |
|---|---|---|---|---|---|---|---|---|---|
| **14 – 21** | | | | | | | | | |
| | Mother´s age | | 0.03 | 0.06 | 0.00 | 0.954 | 1.00 | 0.89 | 1.12 |
| | **Education level** | School* | 17.14 | 2307.71 | 0.00 | 0.994 | 2809873.8 | 0.00 | - |
| | | **High school** | 1.83 | 0.73 | 6.22 | **0.013** | 6.26 | 1.48 | 26.01 |
| | Work | Unemployed | 1.14 | 0.89 | 1.62 | 0.204 | 3.13 | 0.54 | 18.21 |
| | Family composition | Extended | -0.55 | 1.45 | 0.14 | 0.156 | 0.58 | 0.03 | 10.07 |
| | | Single | 0.97 | 0.70 | 1.88 | 0.482 | 2.63 | 0.66 | 10.45 |

| | | | | | | | | | |
|---|---|---|---|---|---|---|---|---|---|
| | Nº children | | 0.25 | 0.47 | 0.29 | 0.589 | 1.28 | 0.51 | 3.22 |
| | Child sex | Female | 0.84 | 0.62 | 1.86 | 0.175 | 2.33 | 0.69 | 7.86 |
| | Night collaboration | No require | -0.64 | 0.77 | 0.68 | 0.414 | 0.52 | 0.11 | 2.41 |
| | | Sometimes | -0.85 | 0.96 | 0.79 | 0.372 | 0.42 | 0.06 | 2.78 |
| | | Always | 1.00 | 0.97 | 1.07 | 0.309 | 2.73 | 0.40 | 18.33 |
| | **Child NW** | | 1.08 | 0.40 | 7.15 | **0.008** | 2.94 | 1.33 | 6.50 |
| | Sleep arrangements | SS | -1.21 | 0.80 | 2.28 | 0.135 | 0.29 | 0.06 | 1.43 |
| | | RS | 0.67 | 1.11 | 0.36 | 0.549 | 1.95 | 0.22 | 17.3 |
| | **Nap** | Always | -0.29 | 1.39 | 0.04 | 0.835 | 0.74 | 0.04 | 11.40 |
| | | Frequently* | 0.98 | 0.00 | - | - | 2.66 | 2.66 | 2.66 |
| | | **Sometimes** | **-1.62** | **0.82** | **3.87** | **0.049** | **0.19** | 0.04 | 0.99 |
| | **Physical activity** | **≥ 4 times/week** | -1.71 | 0.94 | 3.32 | **0.068** | 0.18 | 0.02 | 1.13 |
| | | **2 - 3 times/week** | -1.58 | 0.75 | 4.47 | **0.034** | 0.20 | 0.04 | 0.89 |
| | | 1 time/week | -1.64 | 1.21 | 1.82 | 0.178 | 0.19 | 0.01 | 2.10 |
| **10- 13** | | | | | | | | | |
| | **Mother´s age** | | -0.97 | 0.042 | 5.38 | **0.021** | 0.90 | 0.83 | 0.98 |
| | **Education level** | School* | 16.69 | 2307.7 | 0.00 | 0.994 | 17719764.1 | 0.00 | - |
| | | **High school** | 1.18 | 0.54 | 4.77 | **0.028** | 3.28 | 1.13 | 9.49 |
| | Work | Unemployed | 0.37 | 0.68 | 0.30 | 0.584 | 1.45 | 0.37 | 5.62 |
| | Family composition | Extended | 0.35 | 1.00 | 0.11 | 0.737 | 1.41 | 0.19 | 10.00 |
| | | Single | 0.39 | 0.53 | 0.54 | 0.466 | 1.48 | 0.52 | 4.26 |
| | Nº children | | -0.36 | 0.33 | 1.20 | 0.271 | 0.69 | 0.36 | 1.33 |
| | Child sex | Female | -0.42 | 0.39 | 1.15 | 0.283 | 0.65 | 0.30 | 1.42 |
| | Night collaboration | No require | 0.40 | 0.48 | 0.70 | 0.403 | 1.50 | 0.58 | 3.87 |
| | | Sometimes | -0.63 | 0.67 | 0.86 | 0.351 | 0.53 | 0.14 | 2.01 |
| | | Always | 0.72 | 0.74 | 0.95 | 0.335 | 2.06 | 0.48 | 8.86 |
| | **Child NW** | | 1.12 | 0.30 | 13.37 | **0.000** | 3.07 | 1.68 | 5.61 |
| | **Sleep arrangements** | SS | -0.44 | 0.60 | 0.52 | 0.463 | 0.64 | 0.19 | 2.11 |
| | | RS | -0.85 | 1.03 | 0.66 | 0.416 | 0.43 | 0.05 | 3.26 |
| | **Nap** | Always | -1.81 | 1.40 | 1.64 | 0.196 | 0.16 | 0.01 | 2.59 |
| | | Frequently* | 19.21 | 8158.1 | 0.00 | 0.998 | 22046551.41 | 0.00 | - |

| | | | | | | | | | |
|---|---|---|---|---|---|---|---|---|---|
| | | **Sometimes** | -0.89 | 0.44 | 4.02 | **0.045** | 0.40 | 0.17 | 0.90 |
| | **Physical activity** | ≥ 4 times/week | -0.89 | 0.62 | 2.00 | 0.159 | 0.41 | 0.12 | 1.40 |
| | | **2-3 times/week** | -0.91 | 0.50 | 3.33 | **0.064** | 0.40 | 0.14 | 1.06 |
| | | 1 time/week | 0.23 | 0.62 | 0.14 | 0.703 | 1.26 | 0.37 | 4.28 |
| **6 – 9** | | | | | | | | | |
| | Mother´s age | | -0.05 | 0.04 | 1.30 | 0.251 | 0.95 | 0.87 | 1.03 |
| | **Education level** | School* | 16.56 | 2307.7 | 0.00 | 0.994 | 15679440.70 | 0.00 | - |
| | | **High school** | 1.29 | 0.55 | 5.47 | **0.019** | 3.64 | 1.23 | 10.70 |
| | Work | Unemployed | -0.49 | 0.75 | 0.43 | 0.517 | 0.61 | 0.14 | 2.65 |
| | Family composition | Extended | -0.66 | 1.13 | 0.34 | 0.569 | 0.52 | 0.06 | 4.80 |
| | | Single | 0.44 | 0.54 | 0.66 | 0.424 | 1.55 | 0.54 | 4.43 |
| | Nº children | | 0.26 | 0.32 | 0.68 | 0.405 | 1.30 | 0.69 | 2.47 |
| | Child sex | Female | 0.03 | 0.39 | 0.00 | 0.945 | 1.03 | 0.30 | 1.42 |
| | Night collaboration | No require | -0.19 | 0.49 | 0.16 | 0.689 | 0.82 | 0.47 | 2.25 |
| | | Sometimes | -0.65 | 0.66 | 0.97 | 0.328 | 0.51 | 0.30 | 2.17 |
| | | Always | 1.08 | 0.69 | 2.40 | 0.094 | 2.95 | 0.14 | 1.91 |
| | **Child NW** | | 0.91 | 0.31 | 8.57 | **0.003** | 2.48 | 1.35 | 4.58 |
| | Sleep arrangements | SS | 0.55 | 0.69 | 0.64 | 0.426 | 1.74 | 0.45 | 6.81 |
| | | RS | 1.31 | 1.00 | 1.70 | 0.191 | 3.71 | 0.51 | 26.60 |
| | Nap | Always | 0.23 | 1.17 | 0.03 | 0.845 | 1.26 | 0.12 | 12.60 |
| | | Frequently* | 1.04 | 125446.1 | 0.00 | 1.000 | 2.84 | 0.00 | - |
| | | Sometimes | -0.27 | 0.42 | 0.40 | 0.525 | 0.76 | 0.32 | 1.76 |
| | **Physical activity** | **≥ 4 times/week** | -1.37 | 0.66 | 4.25 | **0.039** | 0.25 | 0.07 | 0.92 |
| | | **2-3 times/week** | -1.05 | 0.50 | 4.29 | **0.038** | 0.35 | 0.13 | 0.94 |
| | | 1 time/week | 0.51 | 0.59 | 0.74 | 0.393 | 1.66 | 0.52 | 5.32 |

Multinomial logistic regression in the group of mothers of school children, showing all variables analyzed. The reference category for the dependent variable was PSQI <5. Variables that had a statistical tendency or were significant are written in bold letters. Reference categories for the independent variables were as follows: for education level, *complete college*; for work, employed; for family composition, *nuclear family*; for child sex, male; for night collaboration, *does not have*; for sleep arrangements, *bed-sharing*; for napping, *never taking naps*; for physical activity (≥ 30 min), *never does*. Subcategories marked with an asterisk (*), the number of mothers was too low for statistical interpretation. Model fit: $R^2$ = 0.39 (Cox & Snell), 0.42 (Nagelkerke); Model $\chi^2$ = 109.2, $p < 0.05$. Abbreviations: B, beta coefficient; CI, confidence interval; NW, night awakening; OR, odds ratio; RS, room sharing; SS, solitary sleep.
